# Optimizing Cognitive Performance Using Memory Enhancing Acute Intermittent Hypoxia (meAIH) in Young Healthy Adults: A Preliminary Study

**DOI:** 10.64898/2025.11.30.25341321

**Authors:** Wendy L. Olsen, Michael Collins, Tiffany S. Jastrzembski, Christopher W. Myers

**Author notes:** Correspondence concerning this article should be addressed to Wendy L. Olsen, Appalachian State University, Department of Rehabilitation Sciences, 1179 State Farm Rd, Boone, NC 28607. The authors have no conflicts of interest to disclose.

## Abstract

The human’s ability to acquire and retain novel information has been investigated via cognitive and physiological approaches. The current study employed a neruoplastic primer, acute intermittent hypoxia (AIH), to elicit beneficial changes in cognitive performance, specifically a declarative memory task, in young, healthy adults. It was hypothesized that participants who were exposed to a single session of a memory-enhancing acute intermittent hypoxia (meAIH; ∼10% oxygen) protocol would demonstrate enhanced performance on a declarative memory task when compared to a SHAM (∼21% oxygen; control) group at an acquisition and one-week retention point. Results indicated that during the initial acquisition phase, the meAIH group performed significantly better on a declarative memory task than the SHAM group (p < 0.05), but not the retention phase. These novel results inform the understanding of cognitive neuroplasticity within young, healthy adults and how meAIH can be used to inform training paradigms for many populations.

## Introduction

The acquisition of new knowledge requires time and resources. Methods for optimizing the acquisition of knowledge and skill often focus on minimizing acquisition times while maintaining knowledge efficacy. Efforts toward this goal can be roughly divided into two different methodological approaches: *cognitive* and *physiological*. Cognitive approaches focus on affecting information processing constraints and mechanisms, while physiological approaches focus on affecting anatomical and chemical processes associated with learning and forgetting. Each approach collectively reflects a fundamental goal: *increasing the rate of knowledge acquisition with better retention.* Methods applying the cognitive approach have demonstrated acquisition and retention benefits using intelligent tutors, personalized training systems, and optimized practice schedules (Jastrzembski et al., 2006, 2017; VanLehn et al., 2011). Although these methods have been shown to improve overall retention, they require distributing repetitions across multiple sessions possessing fewer number of practice opportunities to facilitate slower knowledge acquisition in favor of more stable retention (i.e., the spacing effect; Ebbinghaus, 1885; Walsh et al., 2023).

Researchers have also focused on physiological methods, such as brain stimulation (e.g., transcranial-magnetic-stimulation; Grabner et al., 2015), pharmacological treatments (e.g., methylphenidate; Frank et al., 2014), and exercise (Borror, 2017) to speed-up knowledge acquisition and improve retention. However, they have yet to demonstrate long-term retention without adverse side effects (i.e., financial costs, malaise, etc.). In the current paper, a physiological therapy, demonstrated to mitigate respiratory, physiologic, and cognitive impairment in spinal cord injury, disease, and elderly populations, was applied to young, healthy adults as a potential means to increase the rate of knowledge acquisition. Further, a cognitive model of learning and retention was implemented to facilitate the identification of the affected memory mechanisms.

Previous work has demonstrated that brief, repetitive, and mild presentations of low oxygen, or *acute intermittent hypoxia* (AIH), has elicited functional benefits without demonstrated pathology in people living with spinal cord injury (Navarrete-Opazo and Mitchell, 2014; Sutor et al., 2021) and other neurodegenerative populations (e.g., amyotrophic lateral sclerosis; Sajjadi et al., 2022). The primary driver of these benefits is the increased release of brain derived neurotrophic factor (BDNF) which promotes neurogenesis and neuroplasticity in respiratory, spinal, and memory systems (Agosto-Marlin and Mitchell, 2017; Lao-Peregŕın et al., 2017).

Importantly, BDNF has been identified as a protein associated with learning and memory through its facilitation of neurogenesis and neuroplasticity in the hippocampal region in human and animal models (Mattson et al., 2004; Meng et al., 2020). Thus, the repeated presentation of AIH has revealed cognitive benefits in individuals suffering from cognitive impairment and cognitively aged individuals (Schega et al., 2013; Wang et al., 2020). Results from these studies indicated that following AIH administration, participants experienced improved accuracy on the California verbal test, the digit-span task (short term memory), the number-combination task (reasoning ability), and the d2-test of attention (visual attention). These results demonstrate AIH’s potential to enhance various aspects of cognitive performance.

It remains unclear if AIH will provide similar benefits to young, healthy individuals. Based on evidence of neurogenesis within the hippocampal system and its relationship to the storage, maintenance, and access to declarative information (Khuu et al., 2021; Lavenex & Lavenex, 2013), an empirical behavioral study was conducted to determine if the administration of an AIH protocol would accelerate the acquisition of novel declarative information and improve its retention within healthy adults. It was hypothesized that participants who received the AIH protocol would reduce retrieval errors within a declarative memory task (i.e., paired associate) at a faster rate than participants within a control protocol, SHAM, and that the advantage would persist over a week-long retention period. It was further hypothesized that AIH would provide no performance benefits in a predominantly procedural memory task (i.e., change signal; further detailed hypotheses are provided after task descriptions in the Method section). A previously validated mathematical model of declarative memory dynamics was applied to help identify the hypothetical declarative memory constructs affected by AIH (Walsh et al., 2018a; Walsh et al., 2018b; e.g., memory decay, activation, etc.). In the following sections the Method and Results are provided with a mathematical model of human knowledge acquisition and retention to identify what processes are most likely affected by AIH.

## Method

First the participant sample is described followed by the experimental procedure, including the approach to providing participants reduced oxygen and the descriptions of the experimental tasks. An initial group of 30 participants (15 per group) was determined to be an appropriate sample based on *a priori* methods and was attempted to be acquired (Schega et al., 2013). It is important to note that this study was not preregistered. The following data are reported.

### Participants

The Institutional Review Board of the University of Florida approved the study (IRB# 202300905), and all participants signed informed consent prior to participation. The informed consent procedure followed the guidelines set forth in the Declaration of Helsinki. Seventeen healthy, young adults (7 females, [μ= 26.7 y/o, SD = 7.2] (Female participants identified as/with: 2 Hispanic/Latin American, 1 Asian/Pacific Islander, and 5 Caucasian/White), 10 males [μ= 27.8 y/o, SD = 8.3] (Male participants identified as/with: 3 Hispanic/Latin American and 7 Caucasian/White)) were recruited and randomly selected to receive either an AIH (μ= 26.2 y/o, SD = 6.5) or SHAM (μ= 28.6 y/o, SD = 9.0) protocol. Participants were asked to refrain from caffeine and nicotine products and to avoid physical exercise for six hours prior to study participation. Additionally, any participants who reported respiratory-related illness or symptoms (e.g., excessive cough/sneezing behaviors, fever, etc.) were dismissed from the study).

### Experiment Procedure

All participants provided informed consent on arrival, were administered a COVID-19 screening test, and females were provided a pregnancy test. If participants tested positive for COVID-19 or had a positive pregnancy test, they were excluded from the study. If a negative result from both tests was elicited, those participants were included in the study. An initial 25 participants were recruited for the study. However, five were dismissed from the SHAM group (3 due to positive test results and 2 due to respiratory-illness reports) and 3 from the AIH group (1 due to fever, 1 requested withdraw, and 1 respiratory illness). The resulting sample included 17 participants. Next, participants were randomly assigned to the memory-enhancing acute intermittent hypoxia (meAIH) or SHAM conditions. Baseline measures were gathered for respiratory rate, oxygen saturation, pulse rate, and blood pressure. Following baseline measures, participants received a meAIH or SHAM preconditioning protocol. If assigned to the meAIH group, each participant received interleaved one-minute bouts of meAIH (10.2% oxygen) and room air (approximately 21% oxygen) 15 times (see Protocol Presentation, Figure 1). Similar to the meAIH condition, participants assigned to the SHAM group received 15 interleaved one-minute bouts of normoxia (21% oxygen) and room air (approximately 21% oxygen). The purpose of exposing the SHAM group to the same procedure as the meAIH group is to eliminate any confounds from protocol differences. Each protocol took between 35 and 40 minutes to administer (Navarrete-Opazo and Mitchell, 2014). Participants’ oxygen saturation, pulse rate, blood pressure, and respiratory rate were monitored during the protocol administration. Once participants completed their respective protocols, they were asked to watch a 60-minute National Geographic video of their choosing in order to allow the neurophysiologic mechanisms of meAIH to take effect.

**Figure 1.**
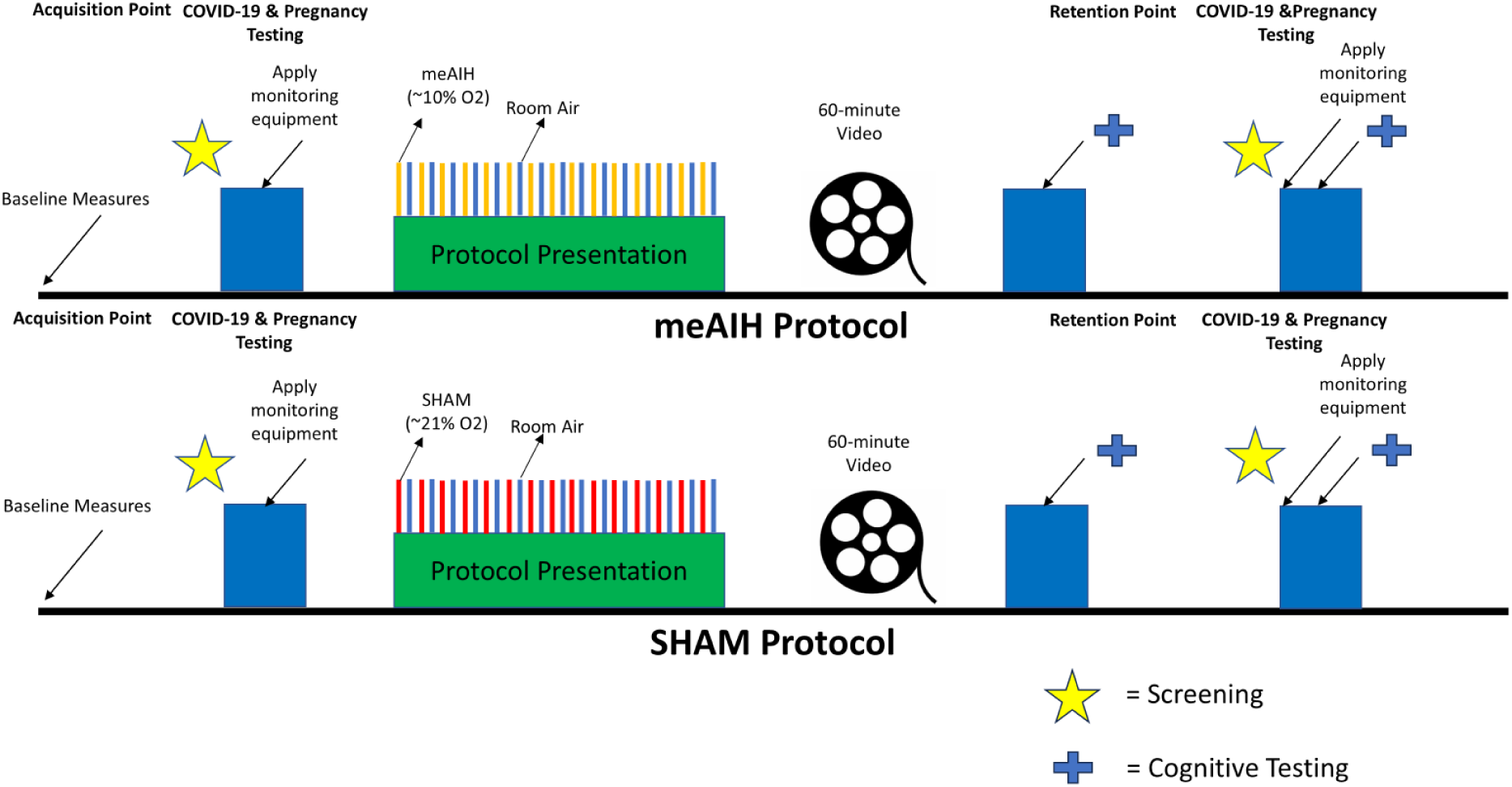
An illustration capturing the study’s schedule in detail. The starting point is the initial screening measures ascertaining the participant’s eligibility (i.e., inclusion/exclusion criteria to the debriefing point (i.e., disclosing their randomized group assignment).

There are at least two broad human memory types: declarative and procedural memory. Declarative memory contains information that can be easily verbalized, such as 2 + 2 = 4 or George Washington was the first president of the United States. Procedural memory contains knowledge composed of procedural steps based on environmental cues and not so easy to verbalize, such as tying shoes or riding a bicycle. To determine if meAIH improved young, healthy adults’ knowledge acquisition, performance on the paired associate task (PAT; Figure 2) was evaluated.The change signal task (CST; Figure 3) was also administered to determine if meAIH affected procedural memory tasks as well as declarative memory tasks. Screen distance was approximately 33 cm for each participant, and all participants had normal or corrected to normal vision.

**Figure 2.**
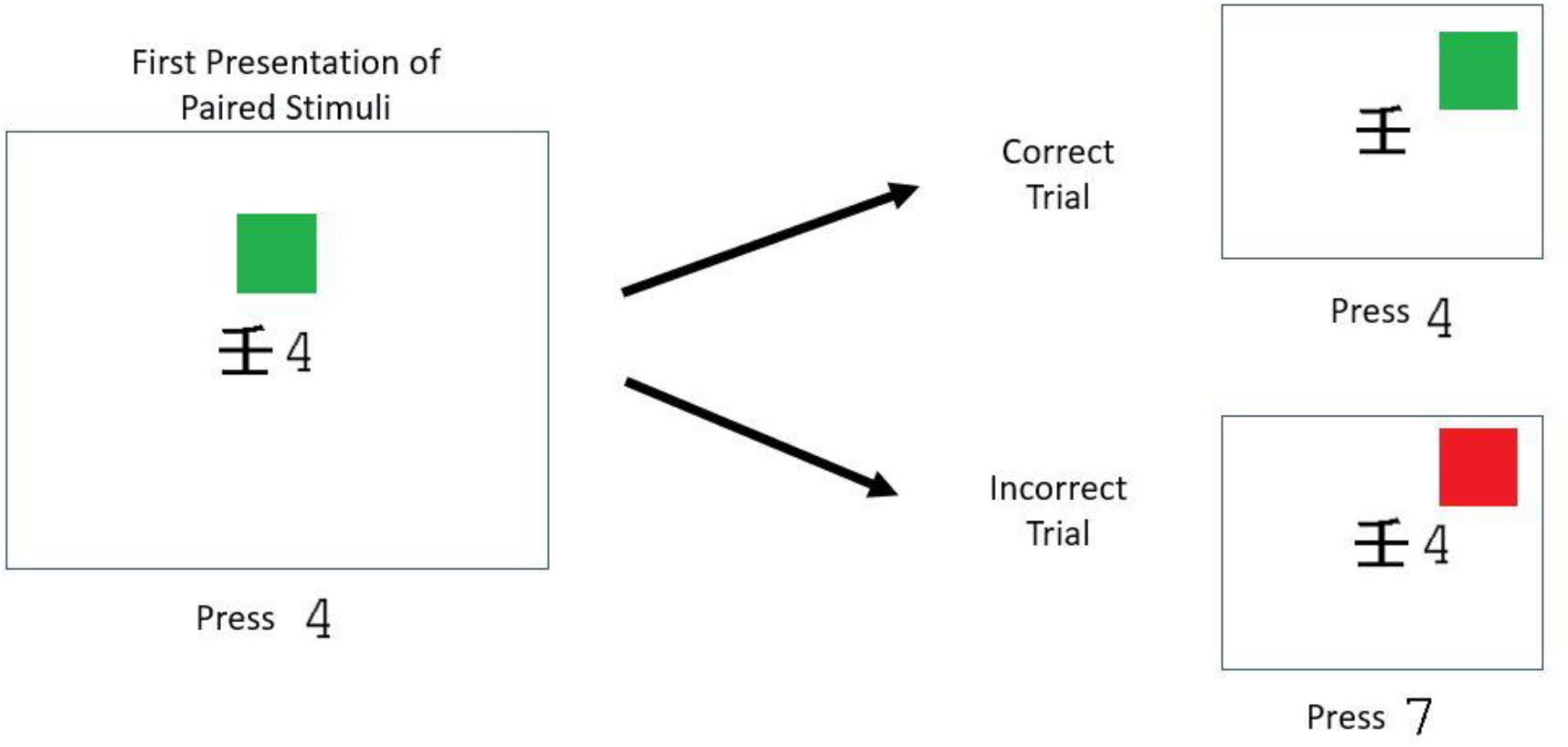
An example of the initial presentation of a stimulus item, a correct trial, and an incorrect trial for the paired associate task (PAT). Participants were provided one training trial. Every proceeding trial, participants were presented with the non-English symbol. If answered correctly, participants received a green square as feedback. If answered incorrectly, participants received a red square and the correct response as feedback.

**Figure 3.**
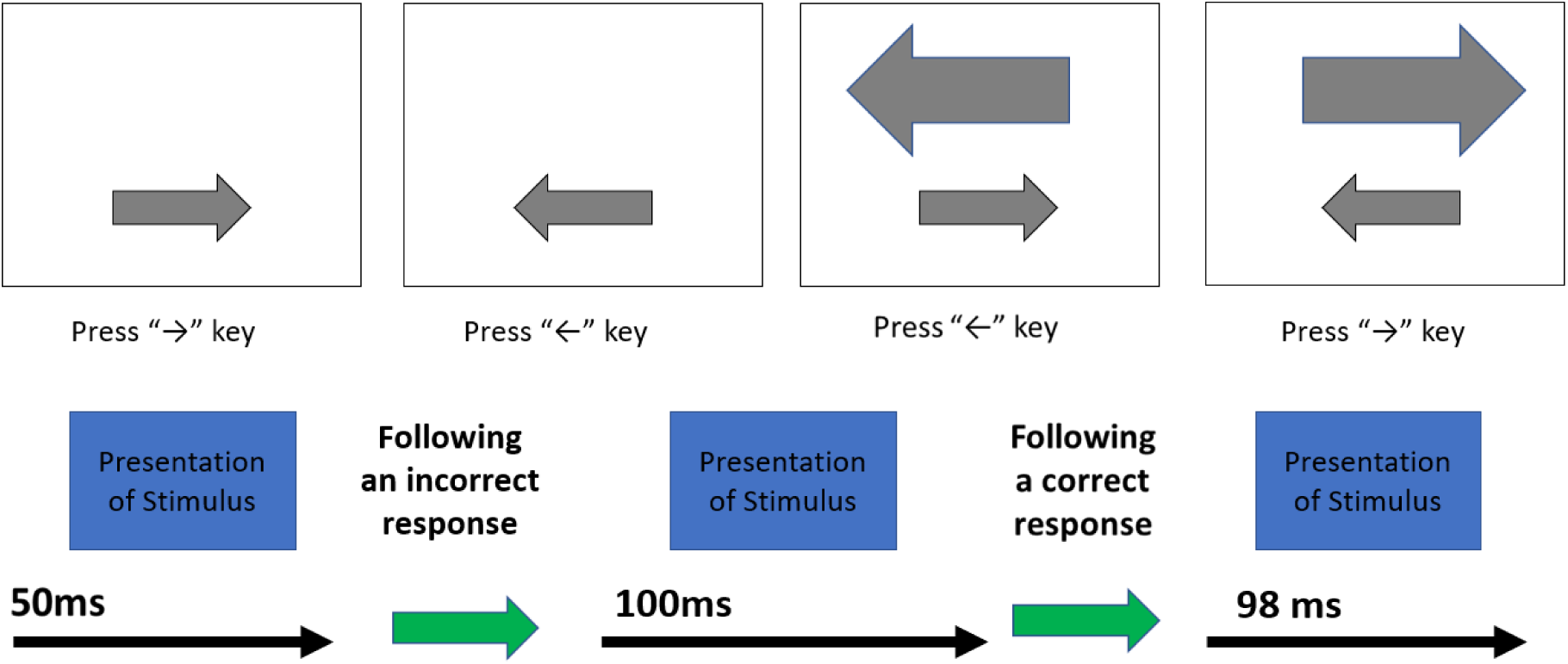
An example of the initial presentation of a stimulus item, a correct trial, and an incorrect trial for the change signal task (CST). Participants are presented with smaller arrows at the bottom of the screen and instructed to respond to the corresponding arrow keys. They are told to change their response to a larger arrow, should it appear at the top of the screen after the smaller arrow appears. If participants answer correctly, the time of the larger arrow appearing increases by 50 msec. If the participants answer incorrectly, the time of the larger arrow appearing decreases by 2 msec.

### Paired Associate Task (PAT)

The paired associate task (PAT) was developed by researchers to understand how humans acquire and maintain connections between two different symbols, or paired associations (e.g., a symbol paired with a number). For the current study, non-English logographs/symbols were paired with Arabic numerals as stimuli and have been used previously to evaluate declarative knowledge acquisition (Halverson and Gunzelmann, 2011).

Stimuli for the PAT consisted of ten black-lined symbols, or non-English logographs, placed on a white background. Each symbol was paired with a number (0-9). Participants were instructed to press the number on the keyboard that was associated with the symbol and received 10 trials for training. The participant was presented with a blank screen for 1000 msec. Next an initial first presentation of a symbol paired with a number was shown; see Figure 2. The stimulus was shown on the screen for a total of 5000 msec. If the participant pressed the correct number associated with the symbol, they received feedback in the form of a green square that was presented for 1500 msec. If the participant pressed the incorrect number, they were provided the feedback with a red square and the correct number for 1500 msec. The PAT consisted of 400 trials in total that were randomly presented in 1 block.

For each participant, response times (RT) and response accuracies (ACC) were recorded. If meAIH produces a benefit to knowledge acquisition, then errors should be eliminated with fewer paired exposures. If the benefit of AIH persists, then there should be fewer response errors in the meAIH group when compared to the SHAM group in the retention phase.

### Change Signal Task (CST)

The change signal task (CST) is an adaptive procedural memory task requiring a participant to respond to a stimulus with a corresponding keystroke (left/right arrow). Stimuli for the CST included a small and large left and right arrow key. Participants were instructed to press the left/right arrow keyboard keys when they saw the small arrow keys appear on the computer screen. Participants were further instructed to inhibit that response if a larger arrow appeared above the smaller arrow and to respond by pressing the arrow key corresponding with the direction of the large arrow (see Figure 3). The participant was presented with a blank screen for 1000 msec. Next, an initial presentation of a right/left small arrow appeared at the bottom of the screen for 1000 msec. On one third of the trials, a larger arrow pointing in the opposite direction of the go signal appeared after a change signal delay (CSD). In this case, subjects were instructed to inhibit their initial response to the go signal (smaller arrow key) and instead respond to the “change signal”, the larger arrow key that appeared. A response ended the trial, which progressed directly to a blank screen and the inter-trial delay. No feedback was presented. If the subject failed to respond within one second after the go signal appeared, the trial timed out.

The CSD was adjusted independently using a step function to maintain a consistent level of difficulty and avoid ceiling effects. Thus, CSDs were constrained to a range of 20 to 800ms, and incorrect responses reduced the CSD by 50ms and correct responses increased the CSD by 2ms; making it harder to inhibit a response. The CST consisted of 300 trials in total that were randomly presented in 1 block. Button response time, total errors made, and accuracy data were collected and analyzed (D’Alberto et al., 2018; see Figure 3). This process requires inhibiting learned reflexes (i.e., inhibits the prepotent motor response for the initial stimulus when followed by a change signal) and is thought to be a marker of impulsivity (Plawecki et al., 2018) and proceduralization.

### Detailed Hypotheses for PAT and CST

Effects of meAIH were hypothesized to affect accuracy performance on PAT. Specifically, a main effect of condition (meAIH | SHAM) on accuracy, where participants receiving meAIH performed with higher accuracy than those receiving the SHAM. Second, a main effect of session (session 1 [acquisition] | session 2 [retention]), where participants receiving meAIH performed with higher accuracy in the acquisition and retention phases versus those participants who received SHAM. Third, a within-session instance x condition interaction as hypothesized, where participants who received meAIH would produce accurate responses at a faster rate within a session (acquisition and retention) when compared to those who received sham. PAT response times were not hypothesized to be affected, only accuracy performance. The presentation of meAIH was not hypothesized to affect performance in the CST.

## Results

Results from the paired associate task are presented first, specifically accuracy followed by response times. Change signal task accuracy and response time results are then provided.

Following the statistical analysis, a mathematical model is introduced and fit to the data to help explain the resulting effects of meAIH. All raw data files, and code, are available in a repository (*see* https://drive.google.com/drive/folders/11Ql1TjQiVdsYm7FQN8543-Pda-EUJHh_).

### Paired Associate Task

To assess the accuracy measures on the PAT, a linear mixed effects (LME) model with a binomial link function using condition (meAIH vs SHAM), session (session 1[acquisition] vs session 2 [retention]), time since an item’s last presentation, session instance, and a quadratic term of session instance to predict accuracy was used. The model also included “participant” as a random effect. The model’s explanatory power was substantial (R^2^ = .46). To ensure that the terms included in the model were necessary, the previously described full model to an intercept-only model (*AIC* = 5859.053) and a model that excluded the quadratic effect of session instance (*AIC* = 4393.753) was compared. Both alternative models were found to have a higher AIC compared to the full model (*AIC* = 4091.692) suggesting the components of the full model were warranted.

The LME model of accuracy found several significant main effects and interactions in line with standard patterns of learning as well as differences between the meAIH and SHAM conditions. The main effects will first be reviewed and then the significant interaction terms. From the significant main effects, three standard learning effects were found to be significant. First, there was a significant positive effect of session (***β*** = 1.28, 95% *CI* [0.80, 1.75], p <.001; Std.***β*** = 0.62, 95% *CI* [0.47, 0.77], effect size = small), with performance in the second experimental session being slightly higher (*M* = .98, *SD* = .14) than the first experimental session (*M* = 0.89, *SD* =.310) (Figure 4 - panel A). Second, there was a significant positive linear effect of session instance on accuracy (***β*** = 1.25, 95% CI [0.89, 1.61], *p* < .001; Std.***β*** = 0.86, 95% CI [0.68, 1.03], effect size - small), with accuracy increasing over the course of a session (Figure 4 - Panel B). Third, there was a significant negative effect of time since an item’s last presentation (***β*** = -0.25, 95% CI [-0.44, -0.06], *p* = 0.011; Std. ***β*** = -0.09, 95% CI [-0.24, 0.05], effect size = small), with accuracy decreasing as the time between presentations of an item pair increased (Figure 4 - Panel C). Finally, there was a significant positive effect of condition on accuracy (***β*** = 1.54, 95% CI [0.67, 2.41], *p* < .001; Std.***β*** = 0.44, 95% CI [0.03, 0.85]), effect size = small), with overall PAT accuracy in the meAIH condition being slightly higher (*M* = .96, *SD* =.20) than the SHAM condition (*M* = .88, *SD* = .31) (Figure 4 - Panel D).

**Figure 4.**
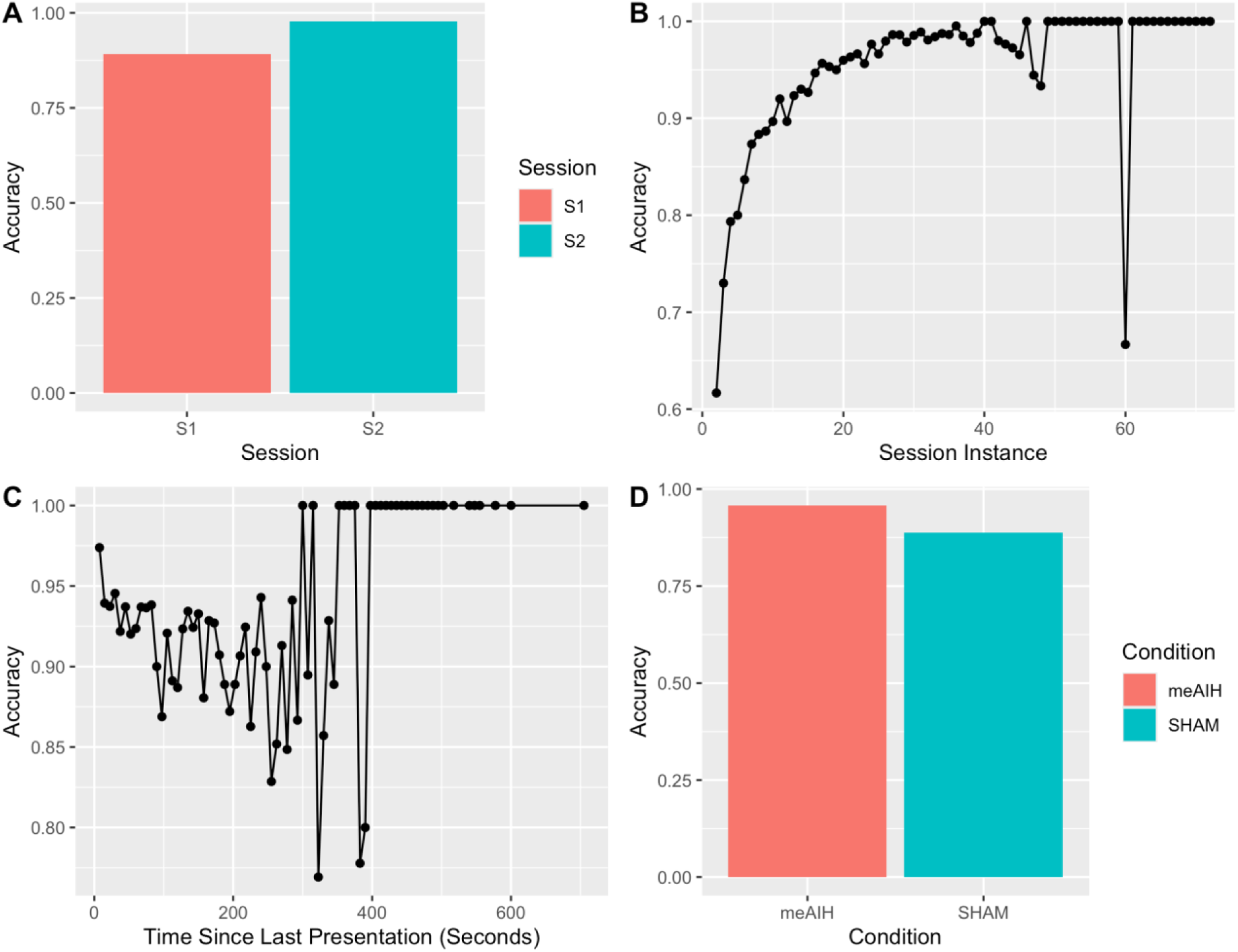
The figure above shows, the average accuracy per session (panel A), the average accuracy per session instance (panel B), the average accuracy per time since the last presentation of an item (panel C), and the average accuracy per condition (panel - D).

Taken together these significant main effects of the different predictor variables of the support for three standard learning effects. The accuracy of participants was higher in the second compared to the first experimental session, suggesting that participants retained information from the first session. Second, the participants’ accuracy improved within a session with each additional presentation of an item pair. Finally, accuracy was found to decrease as the delay between presentations of an item increased, showing small but significant effects of within-session decay. In addition to these standard learning effects, there was a significant overall effect of condition, with performance being higher in the meAIH condition compared to the SHAM condition, suggesting that the meAIH manipulation improved overall stimuli recall.

In regards to the interaction terms, three significant interactions were found. First, there was a significant interaction between session and the linear effect of session instance (***β*** = -1.46, 95% CI [-2.17, -0.75], *p* < .001; Std. ***β*** = -0.39, 95% CI [-0.57, -0.20], effect size = very small) (Figure 5 - Panel A). The linear effect of session instance was found to be greater during session 1 (***β*** = 1.236) compared to session 2 (***β*** = 0.364) (*z.ratio* = 4.802, p < .001). Second, there was a significant interaction between the quadratic effect of session instance and condition, with the trend for each condition being stronger in the meAIH condition (***β*** = -0.499) compared to the SHAM condition (***β*** = -0.150) (*z.ratio* = 2.150, *p* < .0315)(Figure 5 - Panel B). Finally, there was a significant interaction between the quadratic effect of session instance, condition, and session (***β*** = 0.78, 95% CI [0.14, 1.41], *p* = 0.017; Std.***β*** = 0.22, 95% CI [0.04, 0.40]) (Figure 5 - Panel C). During the first session, the trend was greater for the participants in the meAIH (***β*** = -0.8111) compared to the SHAM condition (***β*** = -0.0738) ( z.ratio = 4.358, p <

**Figure 5.**
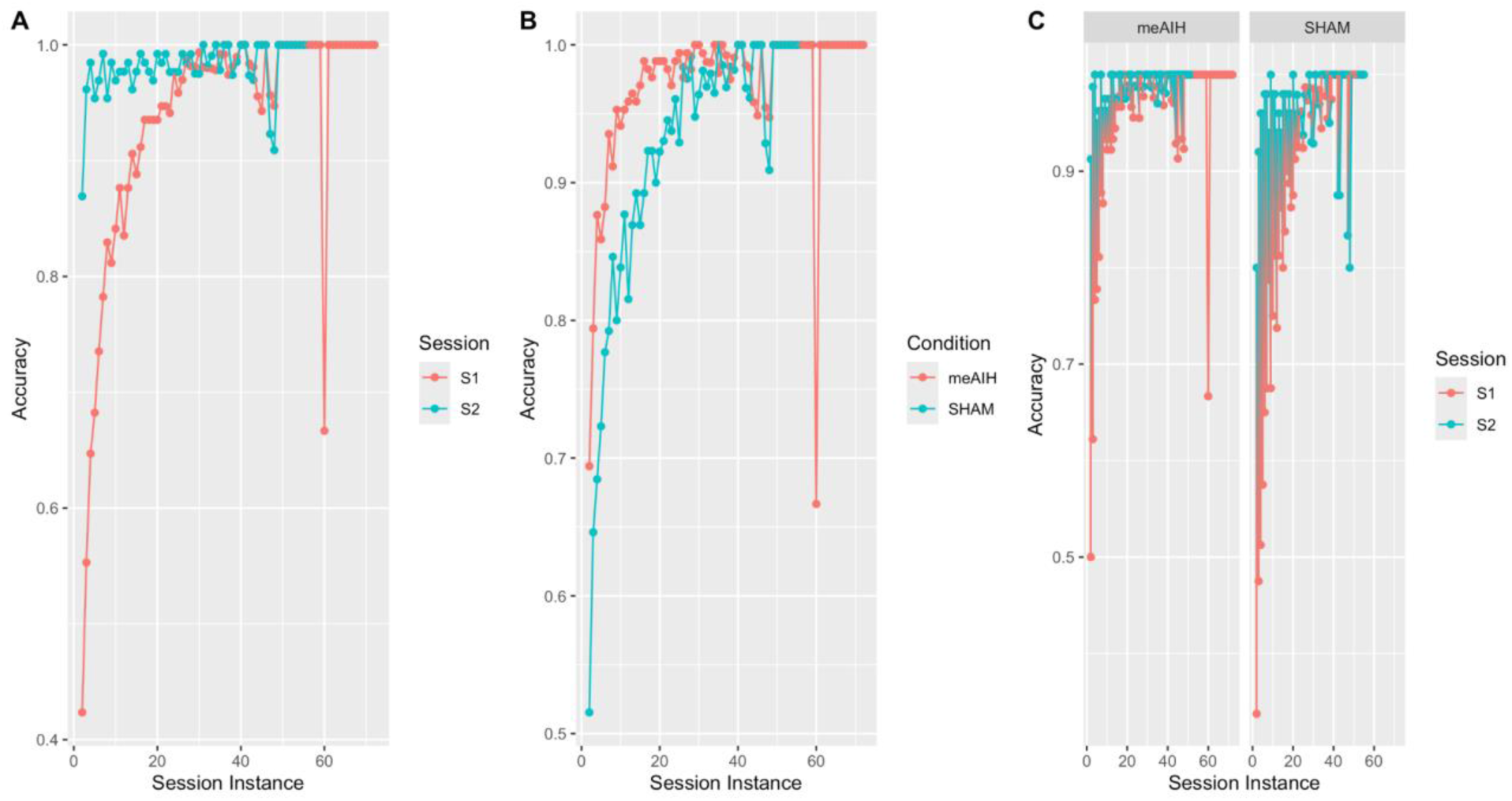
The figure above shows the average accuracy for each session across each session instance (panel A), the average accuracy per session per condition across each session instance (panel B), and the average accuracy for each condition and session per session instance (panel C).

.0001). However, during the second session, no significant difference was found between the trend for the meAIH (***β*** = -0.2256) and SHAM (***β*** = -0.1862) (*p* > .05).

From these significant interactions, accuracy increased at a faster rate in the first compared to the second experimental session. This suggests that the majority of learning took place during the first experimental session. Furthermore, between the meAIH and SHAM condition, accuracy was found to increase at a faster rate during initial trials in the meAIH condition compared to the SHAM condition during the first experimental session. From this, it was concluded that the effect of the meAIH had a greater positive effect on knowledge acquisition during the first experimental session compared to the SHAM condition. However, we failed to find significant evidence of a benefit to retaining the acquired knowledge after one week.

To account for participants’ response times on the PAT a general linear model with a Gamma link function with condition, session, time since last presentation, and both a linear and quadratic effect of session instance as predictors were used. No random effects of participants were included in the model due to the fact that the inter-class correlation (icc), a measure of how much variance in the data can be explained due to the random effects of participants, was very low (icc = .03). The model’s explanatory power was found to be moderate (R^2^ = 0.24). As with the accuracy, we compared the full model to both an intercept-only model (AIC = 16301.8) and a model with only a linear effect of session instance (AIC = 13648.22) finding that the full model had the lowest AIC (*AIC* = 13429.19) compared to both alternative models.

As when investigating accuracy, the GLM model of response time found both significant main effects and interactions. The main effects of the prediction variables on response times will be reviewed first, and then the significant interaction terms. As found with accuracy, there were several significant main effects showing standard learning effects. First, there was a significant positive effect of session on response time (***β*** = 0.26, 95% CI [0.23, 0.28], *t*(11381) = 19.11, *p* < .001; Std. ***β*** = 0.08, 95% CI [0.07, 0.08], effect size: very small) with response times being slightly slower in the second experimental session (*M* = 1.17, *SD* = 0.416) compared to the first session (*M* = 1.47, *SD* = 0.719) (Figure 6 - panel A). Second, a negative effect of the time since an item’s last presentation (***β*** = -0.04, 95% CI [-0.05, -0.03], t(11381) = -6.53, p < .001; Std.***β*** = -0.03, 95% CI [-0.03, -0.02]), with response times increasing as the delay between presentations of an item increased (Figure 6 - Panel B) was detected. Finally, there was a significant effect of both linear (***β*** = 0.08, 95% CI [0.06, 0.10], t(11381) = 7.39, p < .001; Std. ***β*** = 0.05, 95% CI [0.04, 0.06]) and quadratic (***β*** = -0.03, 95% CI [-0.04, -0.02], t(11381) = -5.75, p < .001; Std. ***β*** = -0.05, 95% CI [-0.06, -0.04]) effect of session instance on response times decreasing with additional presentations of an item within a session (Figure 6 - Panel C). In addition to the standard learning effects, there was a significant positive effect of condition on response time (***β*** = 0.15, 95% CI [0.13, 0.17], *t*(11381) = 15.45, p < .001; Std. ***β*** = 0.04, 95% CI [0.03, 0.04], effect size: very small ), with response time being slightly slower in the peIAH (*M* = 1.25, *SD* = 0.521) meAIH group when compared to the SHAM (*M* = 1.47, *SD* = 0.737) condition (Figure 6 - Panel D).

**Figure 6.**
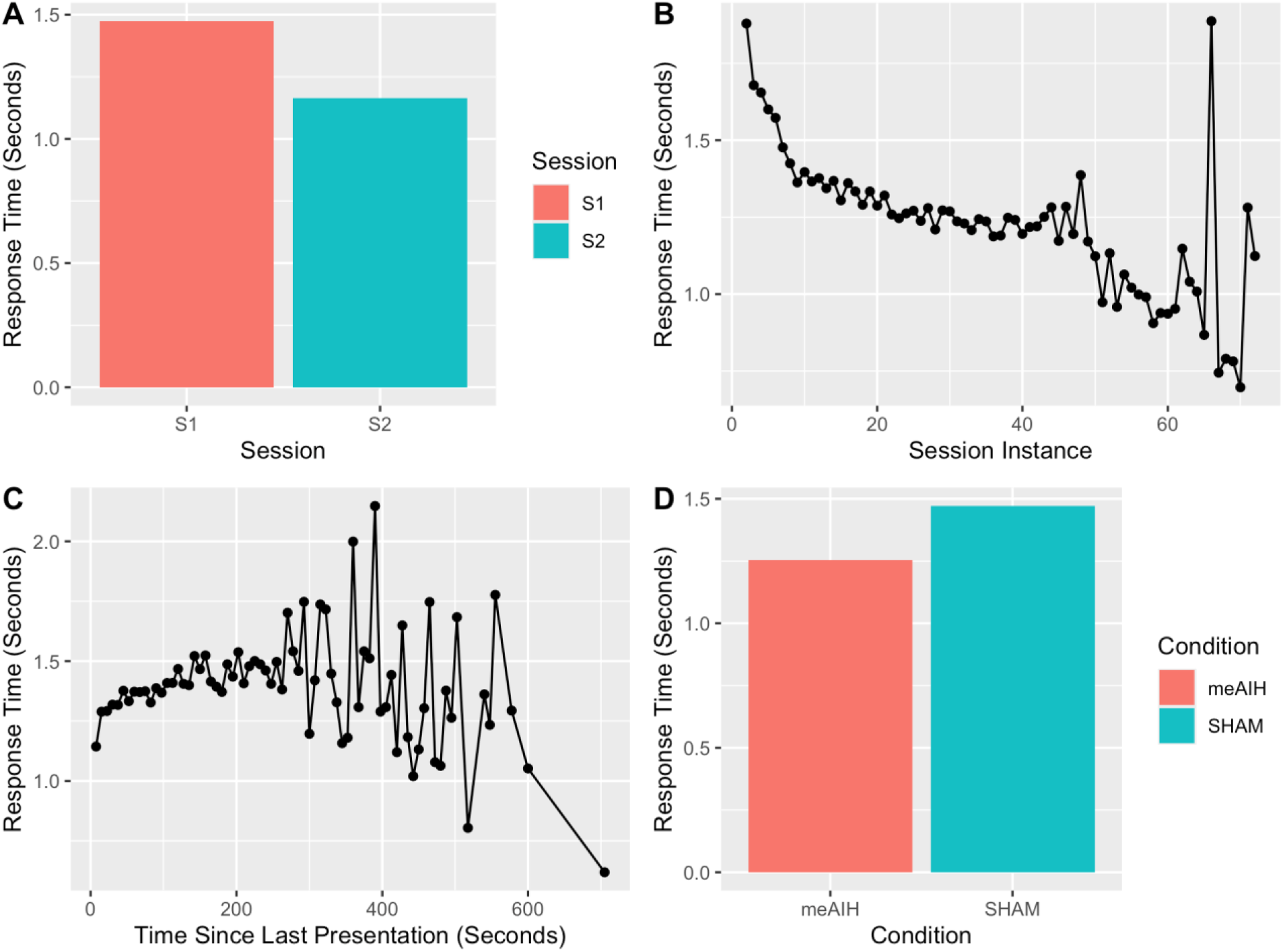
The figure above shows, the average response time per session (panel A), the average response time per session instance (panel B), the average response time per time since the last presentation of an item (panel C), and the average response time per condition (Panel D).

In addition to the main effects there were three significant interactions between the predictor variables and response times. First, there was a significant two-way interaction between condition and session (***β*** = -0.15, 95% CI [-0.19, -0.12], *t*(11381) = -8.43, *p* < .001; Std.***β*** = -0.04, 95% CI [-0.04, -0.03], effect size: very small) (Figure 7 - Panel A). During the first session, response times were slightly faster in the meAIH (*M* = 1.32, *SE =* 0.00889) compared to the SHAM (*M* = 1.61, *SE* = 0.01230) conditions (*t*(11381) = -19.751 *p* <.0001). During the second session a significant effect between the response times of the meAIH (M = 1.17 *SE* = 0.00840) and SHAM (*M* = 1.16, *SE* = 0.01150) conditions (*p* > .05) was not detected (Figure 7 - panel A). Second, there was a significant two-way interaction between the linear effect of session instance and session (***β*** = -0.06, 95% CI [-0.10, -0.02], t(11381) = -3.00, *p* = 0.003; Std. ***β*** = -0.03, 95% CI [-0.04, -0.03], effect size: very small) (Figure 7 - Panel B). The linear effect of session instance was larger in the first session (***β*** = 1.236, SE.***β*** = 0.0871) compared to the second session (***β*** = 0.364 se.***β*** 0.1600) (z.ratio = 4.802, *p* <.0001) (Figure 7 - panel B). Finally, there was a significant two-way interaction between condition and quadratic effect of session instance (***β*** = -0.02, 95% CI [-0.03, -2.35e-03], t(11381) = -2.29, *p* = 0.022; Std. ***β*** = -7.87e-03, 95% CI [-0.02, -6.49e-04], effect size very small) (Figure 7 - Panel C). The trend in response times was greater in the meAIH condition (***β*** = -0.499, SE.***β*** =0.109) compared to the SHAM condition (***β*** = 0.150, SE.***β*** = 0.120) (z.ratio = 2.150, p < 0.0315) (Figure 7 - panel C).

**Figure 7.**
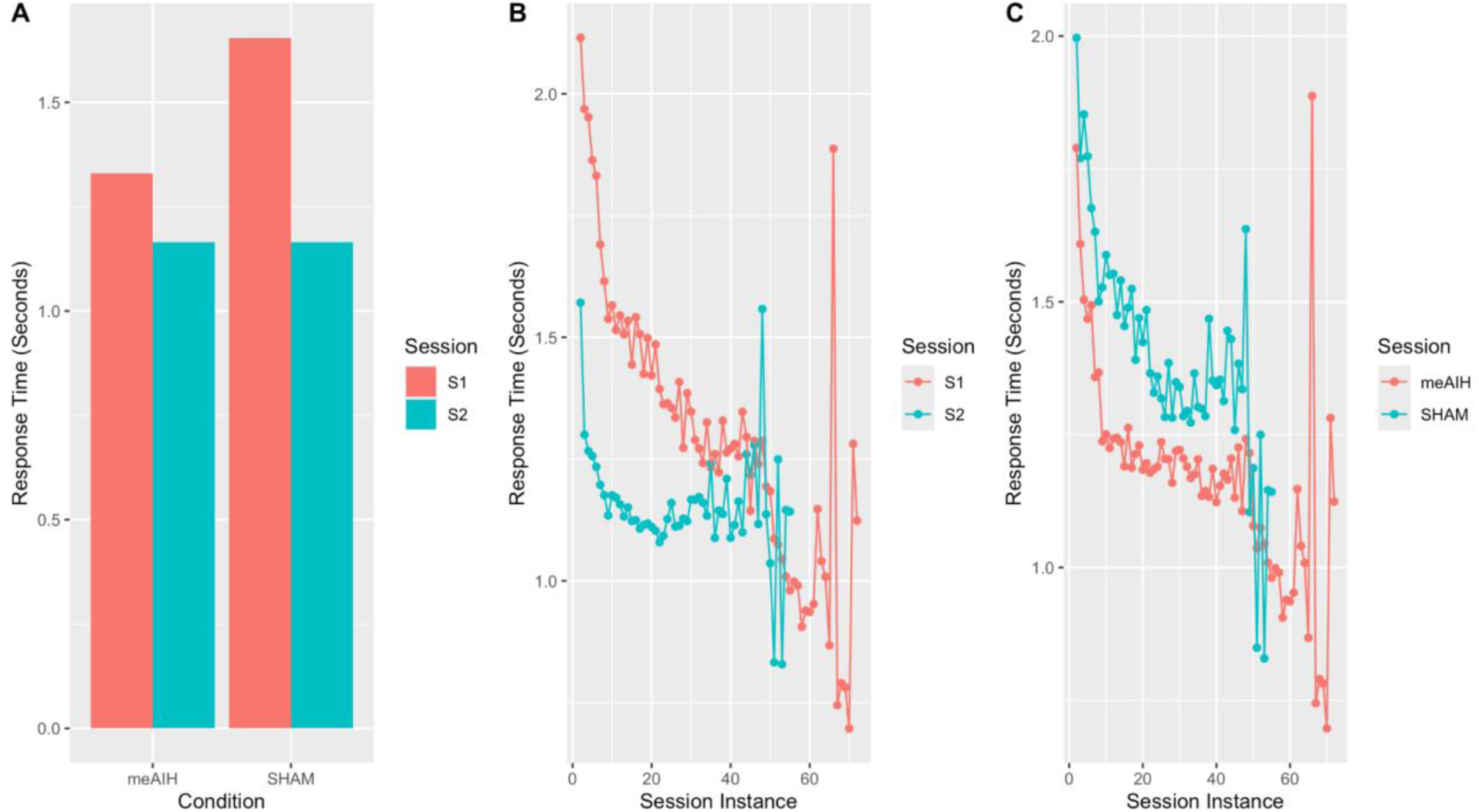
The figure above shows the average response time for each session and condition (panel A), the average response time per session per session (panel B), and the average response time for each condition across session instances (panel C).

Taken together these results of response time on the PAT complement the effects found for accuracy. As with accuracy, the participants’ response times followed the standard learning effects of session, session instance, and time since an item’s last presentation were found, showing the expected learning effects. Additionally, a main effect of the condition was found with participants in the meAIH condition having an overall faster response time compared to participants in the SHAM conditions. Furthermore, again there were significant interactions between response times and session instances, being greater in the first compared to the second condition. Finally, response times during the first experimental session response times were faster in the meAIH condition compared to the SHAM condition as accuracy increased.

Consequently, we can rule out a speed-accuracy tradeoff as an explanation for improved accuracy: participants are not slowing their responses to ensure correct response.

### Change Signal Task

To account for participants’ accuracy on the CST a logistic mixed effects model was used to predict accuracy with condition (meAIH vs. SHAM), session (session 1 and session 2), trial type (congruent - small and big arrow facing the same direction, incongruent trial - small and big arrow facing opposite directions), and change signal delay (CSD). The model included a “participant” variable as a random effect. The model’s total explanatory power is substantial (conditional R^2^ = 0.57) and the part related to the fixed effects alone (marginal R^2^ = .27).

The model found only two significant main effects. First, there was a significant main effect of condition (***β*** = 2.45, 95% CI [0.83, 4.08], *p* < 0.003; Std. ***β*** = 0.23, 95% CI [-0.54, 1.00]) (Figure 8 - panel A), with accuracy being slightly lower in the meAIH (M = 0.925, SD = 0.263 ) compared the SHAM condition (M = 0.964 SD = 0.186)Second, there was also a negative main effect of CSD (***β*** = -9.25, 95% CI [-14.55, -3.96], p < .001; Std. ***β*** = -0.09, 95% CI [-0.80, 0.61]), with accuracy decreasing as the CSD increases (Figure 8 - Panel B). The main effect of meAIH producing lower accuracy was a surprise; however, the main effect of accuracy decreasing with CSD increasing is a typical result from the CST.

**Figure 8.**
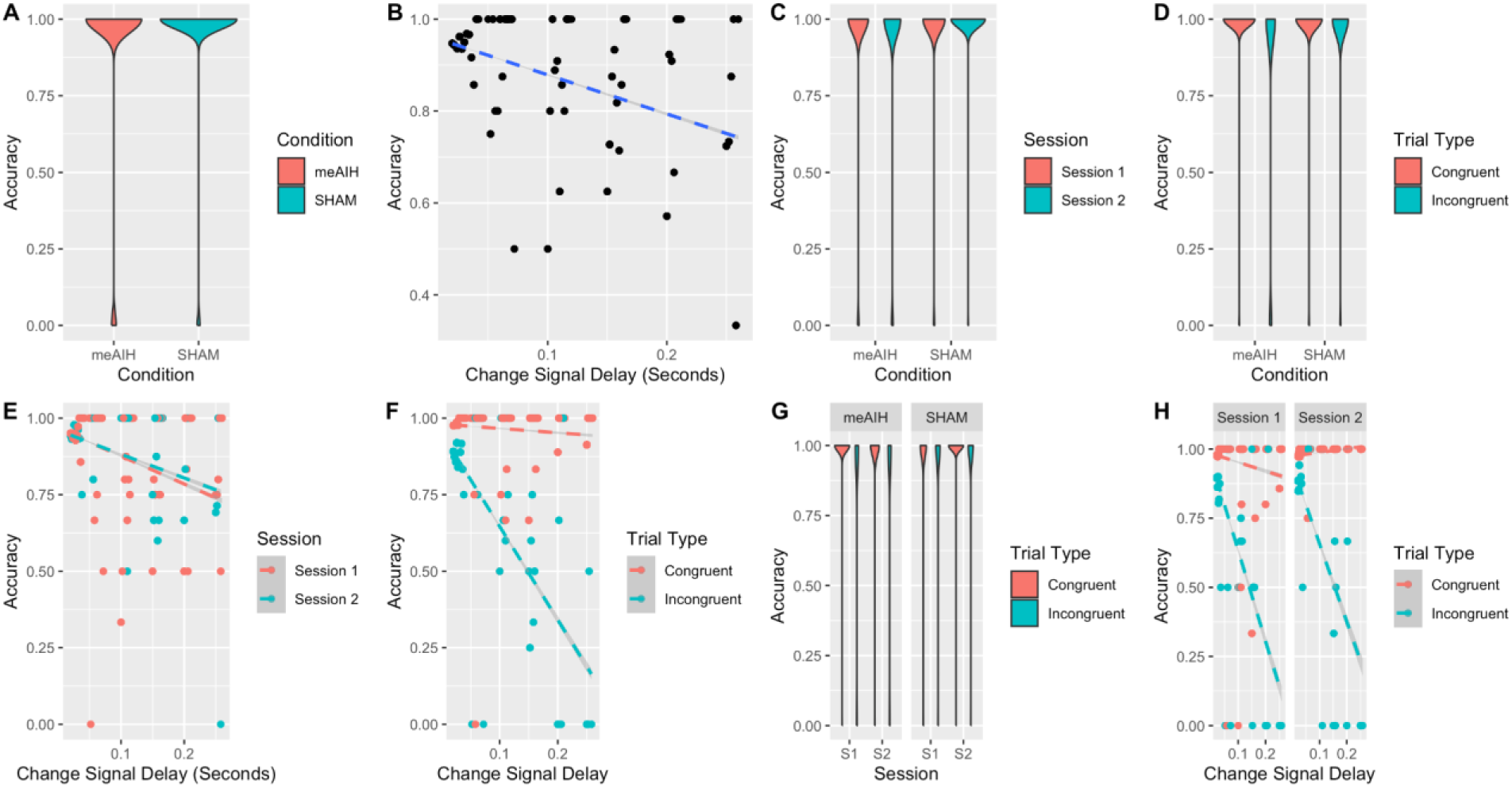
The figure shows the significant the main effects and interactions found in accuracy on the CST task.

Along with the main effects, there were six significant interactions. First, there was a significant negative interaction between condition and session (***β*** = -2.59, 95% CI [-3.67, - 1.51], *p* <.001; Std. ***β*** = -0.48, 95% CI [-0.72, -0.24], effect size: small) (Figure 8 - Panel C). A post-hoc test found that there were two significant differences. First, there was a significant decrease in accuracy on the CST in the meAIH condition between the first session (*M* = 0.936, SD = 0.245) and second (*M* = 0.913, SD = 0.282) session (z.ratio = -3.853, *p* < .001). Second, there was a difference in accuracy on the CST in the SHAM condition during the first (*M* = 0.951, SD = 0.215) and second (M =0.982, SD = 0.133) session (z.ratio = 2.954, *p* < .001). No significant differences were found between the meAIH and SHAM conditions on either the first (*p* > .05) or second session (*p* > .05).

Second, there was a significant negative interaction between condition and trial type (***β*** = -3.45, 95% *CI* [-4.22, -2.67], *p* < .001; Std.***β*** = -0.52, 95% *CI* [-0.69, -0.34]). A post-hoc test revealed several significant differences (Figure 8 - Panel D). First, there was a significant difference between trial type and in the SHAM (z.ratio = 3.09, p < .001) and meAIH (z.ratio = 24.88, p < .001) condition, with accuracy being higher on congruent trials (SHAM: *M* = 0.973 *SD* = 0.162; meAIH: *M* = 0.979 *SD* = 0.144) compared to the incongruent trials (SHAM: *M* = 0.821 *SD* = 0.383; meAIH: *M* =0.821, *SD* = 0.383).

Third, there was a significant interaction between session and CSD (***β*** = 39.92, 95% CI [22.04, 57.80], *p* < .001; Std. ***β*** = 0.35, 95% CI [-0.43, 1.13], effect size: very small) (Figure 8 - Panel E). The trend in the CSD was found to be smaller in the second (***β*** = 2.99 SE = 4.75) compared to the first session (***β*** = 17.90 SE = 2.03) (z.ratio = -20.9, p < .01). Fourth, there was a significant interaction between trial type and CSD (***β*** = -20.85, 95% CI [-30.88, -10.82], *p* < .001; Std.***β*** = -0.40, 95% CI [-0.91, 0.10]) (Figure 8 - Panel F). A post-hoc test revealed a significant difference in the trend line between different CSD values for congruent (***β*** = 9.22, SE = 4.76) and incongruent (***β*** = -24.13, SE = 2.24) trials.

Fifth, there is a significant three-way interaction between condition, session, and trial type (***β*** = 2.29, 95% CI [1.00, 3.57], *p* < .001; Std. ***β*** = 0.28, 95% CI [0.10, 0.46], effect size: very small) (Figure 8 - Panel G). For congruent trials during the first session a significant difference between the accuracy on the CST was found, with accuracy being slightly higher in the meAIH condition (*M* = 0.988, *SD*= 0.107) compared to the SHAM (*M* = 0.958, *SD*= 0.200) condition (z.ratio = 0.0848, *p* < .04). Furthermore, during the first session for the both the meAIH (z.ratio = 39.5118, p < .001) and SHAM (z.ratio = 6.5025, p < .0001) condition, there was a significant difference between the accuracy on both the congruent and incongruent trials (meAIH: *M* = 0.829, *SD* = 0.376; SHAM: *M* = 0.958, *SD* = 0.200). Looking between the first and second session there was a significant difference between the accuracy on consistent trials in the meAIH condition decreasing from the first to the second (*M* = 0.968, *SD*= 0.177) condition (z.ratio = 2.5736, p < .01).

Finally, there was a significant negative three-way interaction between session, trial type, and CSD (***β*** = -33.26, 95% CI [-51.86, -14.65], p < .001; Std. beta***β*** = -0.20, 95% CI [-0.76, 0.36]) (Figure 8 - Panel H). A post hoc test revealed that between the first and second session there was a significant difference between the trend between CSD for congruent (session 1: *β* = -8.95, SE = 2.48, session 2: *β* = 27.40 SE = 8.71) trials (z ratio = -36.36).

To account for the response time in the CST task a general linear mixed model, with a gamma link function was used to predict response times using condition, session, and trial type as predictors. The model also included a random effect of “participants”. The model’s total explanatory power is substantial (R^2^ = 0.66) and the part related to the fixed effects alone (marginal R^2^ = .59).

All three of the predictor variables were found to have a significant effect on response times in the CST task. First, there was a statistically significant and positive main effect of condition (*β* = 0.63, 95% CI [0.33, 0.93], *t*(8527) = 4.09, *p* < .001; Std. *β* = 0.17, 95% CI [0.02, 0.32]), with the response times on the CST being faster in the meAIH condition (*M* = 0.390, *SD* = 0.0915) compared to the SHAM condition (*M* = 0.469, *SD* = 0.146) ( z ratio = -0.296 , *p* < .05) (Figure 9 - panel A). Second, there was a significant effect of session on response times (*β* = 0.58, 95% CI [0.53, 0.62], *t*(8527) = 25.63, *p* < .001; Std. *β* = 0.12, 95% CI [0.11, 0.13]), with response times on the CST being slightly slower on the first (*M* = 0.448, *SD*= 0.143) compared to the second experimental condition ( *M* = 0.391, *SD*=0.0831) (Figure 9 - panel B). Third, there was a significant statistical effect of trial type on response times in the CST (*β* = -0.18, 95% CI [-0.22, -0.15], *t*(8527) = -10.42, *p* < .001; Std. *β* = -0.15, 95% CI [-0.16, -0.14]), with response times on the CST being slightly faster on congruent (*M* = 0.403, *SD* = 0.123) trials when compared to incongruent ( *M* = 0.459, *SD* = 0.116 ) trials (Figure 9 - panel C).

**Figure 9.**
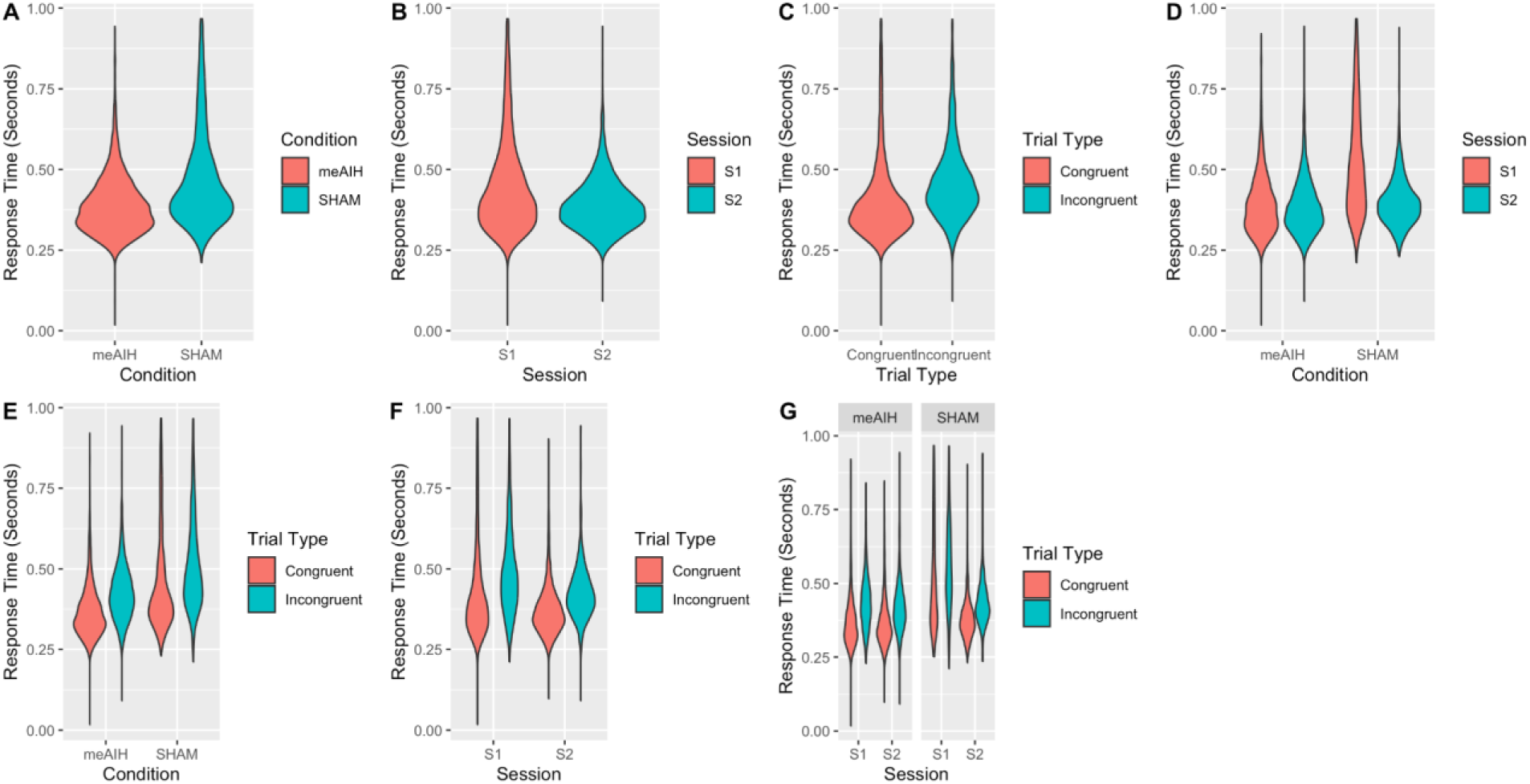
The significant main and significant effects on the response times on the CST task.

Along with the main effects, there were four significant interactions. First, there was a significant two-way interaction between condition and session (*β* = -0.55, 95% CI [-0.61, - 0.50], *t*(8527) = -18.53, *p* < .001; Std. *β* = -0.12, 95% CI [-0.14, -0.11]) (Figure 9 - panel D). A post-hoc test revealed that there was a significant difference in response times on the CST during the first (M = 0.520, *SD* = 0.215) and second (*M* = 0.399, *SD* = 0.133) session in the SHAM condition (z.ratio = -26.947, *p* <.0001). Second, there was a significant two-way interaction between condition and trial type on response times in the CST (*β* = -0.18, 95% CI [-0.24, -0.13], t(8527) = -6.81, *p* < .001; Std. *β* = -0.03, 95% CI [-0.04, -0.02]) (Figure 9 – panel E). A post-hoc test revealed a significant difference between the response times on the CST during congruent (meAIH: *M* = 0.370 *SD* = 0.144; SHAM: *M* =0.450, *SD* = 0.162) and incongruent (meAIH: *M* =0.428, *SD* = 0.383; SHAM: *M* =0.504, *SD* = 0.225) trials in both the meAIH and SHAM condition (meAIH: z.ratio = 23.846, *p* <.0001, SHAM: z.ratio = 15.183, *p* <.0001).

Third, there was a significant two-way interaction between session and trial type (*β* = - 0.12, 95% CI [-0.18, -0.06], t(8527) = -3.74, p < .001; Std. *β* = -9.35e-03, 95% CI [-0.02, 8.59e-04]) (Figure 9 - panel F). A post hoc test revealed that between sessions there was a significant difference between the response times on the CST between the first and second session for both congruent and incongruent trials (z.ratio = -20.473, *p* <.0001, z.ratio = -13.775, *p* <.0001). For both congruent (Session 1: *M* = 0.428 SD = 0.145, Session 2: *M* = 0.373 SD = 0.0784) and incongruent (Session 1: *M* = 0.488 *SD* = 0.132, Session 2: *M* = 0.425, *SD* = 0.0810) trials response times on the CST were found to decrease slightly.

Finally, there was a significant three-way interaction between condition, session, and trial type (*β* = 0.14, 95% *CI* [0.05, 0.22], *t*(8527) = 3.11, *p* = 0.002; Std. *β* = 0.02, 95% CI [5.90e-03, 0.03]) (Figure 9 - panel G). A post-hoc test revealed that there was a significant difference between the response time on the CST task on incongruent trials in the SHAM condition between the first and second session (z.ratio = -16.70, *p* < .001), with response times being slightly slower in the first session (*M* = 0.558, *SD* = 0.145) compared to the second session (*M* = 0.432, *SD* = 0.0705).

## Results Summary

To quickly summarize the results, meAIH produced effects supporting the knowledge acquisition hypotheses. Specifically, there was a main effect of condition (meAIH | SHAM) on accuracy, where participants receiving meAIH performed with higher accuracy than those receiving the SHAM. Second, there was a main effect of session (session 1 [acquisition] | session 2 [retention]), where participants receiving meAIH performed with higher accuracy in the acquisition and retention phases versus those participants who received SHAM. Third, there was a within-session instance x condition interaction, where participants who received meAIH produced accurate responses at a faster rate within a session (acquisition and retention) when compared to those who received SHAM. Interestingly and surprisingly, there were also significant decreases in response times between the meAIH and SHAM conditions, between sessions, and PAT response times in the meAIH condition decreased at a faster rate than response times from participants receiving SHAM.

There were no effects of meAIH hypothesized to occur in the CST. However, meAIH had a negative effect on accuracy compared to SHAM as well as decreasing response times relative to SHAM.

### Mathematical Modeling of Knowledge Acquisition & Retention under meAIH

To evaluate the learning performance of participants on the PAT and infer mechanistic differences in the learning between the meAIH and SHAM conditions, multiple hierarchical implementations of a well-supported mathematical model of knowledge acquisition and retention were fit to participants’ PAT performance. In the current section the general model is described and applied to the data and results from the PAT. A model comparison is provided to help better understand how meAIH affects declarative memory processes.

### The Predictive Performance Equation

The predictive performance equation (PPE) was selected to model the relevant cognitive constructs of knowledge acquisition and retention in the experiment due to its history of successfully accounting for the dynamics of knowledge acquisition and retention (Jastrzembski et al., 2006; Walsh et al., 2018a). The PPE is a mathematical model of learning and retention, which accounts for three different memory phenomena. The first is the power law of learning, performance improves as a function of practice (Newell and Rosenbloom, 2013). Second, the power law of forgetting, performance decreases as a function of the time in between instances of practice. Third, the spacing effect, when practice opportunities are distributed over time (spaced schedule) performance improves at a slower rate but can be retained for a longer duration compared to when practice opportunities occur in more recent succession (i.e., massed practice schedule; Stocco et al., 2024). The PPE is composed of seven different equations which are used to account for the participant’s performance (Accuracy [ACC] and response time [RT]). The core of PPE is the model’s activation term (M_i_, 1), which represents the strength of an item (i) in memory. An item’s activation is a product of the learning and decay term, which are determined by the individual’s learning schedule and estimated learning parameters.

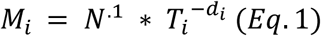

The learning term (N^.1^) is determined by the number of exposures an individual has had with a particular item (N) raised to a constant learning rate of .1^1^. The learning term accounts for the power law of learning. The forgetting term 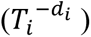 is determined by the model time (*T_i_*, see equation 2) and the decay rate (*d_i_*, 4) and accounts for the power law of learning and spacing effect. Model time is the weighted sum of an individual’s learning schedule (see equations 2 and 3).

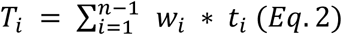

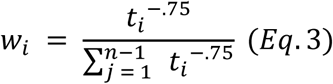

The decay parameter (4) is determined by two free parameters *b, m, a,* and the stability term (ST_i_, 5). The two free parameters control the amount of memory decay between instances of practice. The stability term is a cumulative average of the lags (lag_j_b j) between instances of exposures (5).

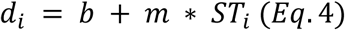

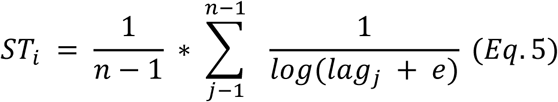

Once the activation term has been determined, it can be transformed into a behavioral measure to account for an individual’s accuracy (ACC_i_) and response time (RT_i_).

To estimate the probability of successfully retrieving an item from memory (ACC_i_ 6), PPE’s activation term is nested within a logistic function, manipulated by the two free parameters (*τ*, *s*) (Walsh et al., 2018a; Walsh et al., 2018b). To estimate the time it takes to recall an item from memory (Response time - Rt_I_ 7) the activation term is nested within an exponential function, manipulated by two free parameters (f, F).

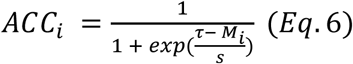

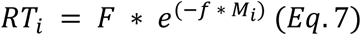

### Applying the PPE

The participants’ accuracy and response time measures from the PAT assessment were fit using three different Bayesian implementations for the PPE (previously discussed), two hierarchical (Figure 10 - left and center pane) and one non-hierarchical (Figure 10 - right pane). Each of the Bayesian implementations is represented as a directed analytics graph (DAG), representing variable types, observed/unobserved, deterministic/stochastic, and model redundancies. All continuous variables are represented by circles, and discrete variables are represented as squares. Observed variables are shaded, while unobserved variables are unshaded. Furthermore, deterministic variables are represented as a single circle while stochastic variables are represented as double circles. Multiple panes within the DAG graph represent redundancies within the model, such as trials, participants, or conditions.

**Figure 10.**
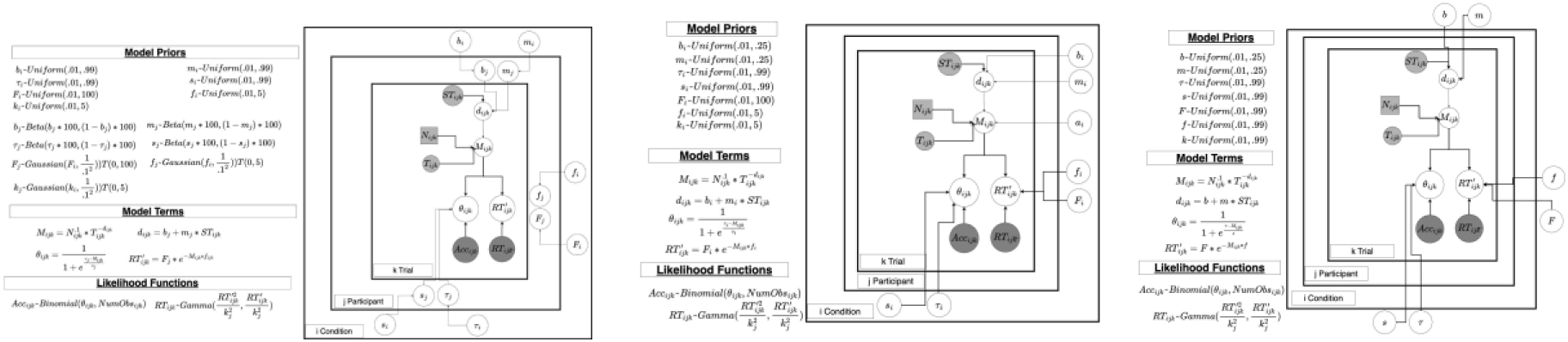
The directed analytic graphs (DAG) for the individual (left), condition (middle), and Null (right) model.

Across all three models, each share the same core PPE model components and likelihood functions, differing only in the relationship between their free parameters. The null model (Figure - right panel) estimates a single set of parameters (b, m, s, tau, F, f, k) across all participants in both the meAIH and SHAM condition. The condition model (Figure 10 - center panel) extends the framework of the Null model and estimates condition specific PPE parameters independently for both the meAIH and SHAM condition (b_i_,m_i_ tau_i_, s_i_,F_i_ f_i_, k_i_). Finally, the individual model extends the framework from of the condition model and estimated condition specific parameters (b_i_,m_i_ tau_i_, s_i_, F_i_ f_i_, k_i_) that then influence the individual parameter estimates for each individual in each condition (b_ij_, m_ij_, tau_ij_, s_ij_, F_ij_, f_ij_, k_ij_). In each of these models the free parameters were sampled from their respective prior distributions and combined with PPE’s terms for each participant thus enabling predictions for both the participant’s accuracy and response times, which are then assessed using either a binomial (accuracy) and gamma (response time) likelihood function.

Finally, all three models were implemented in JAGS (Plummer 2003) and were run for 20000 iterations, with a 10000-sample burn-in period implemented across three independent Markov chains. To ensure that each model converges, all models were checked to ensure that they had an R-hat (Gelman & Rubin, 1992) metric of less than 1.1.

Differences in the participant’s learning performance on the PAT, between the meAIH and SHAM condition, were conducted using a model comparison comparing three different hierarchical implementations of the PPE. A comparison of multiple hierarchical structures allows for the evaluation of the relationship between the PPE’s parameters between the meAIH and SHAM condition. From the model selection we used the most parsimonious model, and we evaluated the differences in the parameters estimated for the meAIH and SHAM conditions. An examination of differences in parameter estimates allows for potential inferences in the mechanisms for the performance differences between the two experimental conditions.

### Model Comparison

To determine the best model to fit the participants’ performance, three different hierarchical implementations of the PPE were compared. The first model was a Null model (Null Model, Figure 10 - left panel), which assumed that there are no differences in the parameters estimated between the meAIH and SHAM condition. The second model was a condition model (condition model, Figure 10 - middle panel) which assumed that there are differences between the parameters estimated for the meAIH and SHAM condition but no individual differences across participants. The third model was an Individual model (Figure 10 - right panel) which assumed that there are differences in the parameters estimated between both the SHAM and meAIH condition and individual participants. To compare these different implementations of the PPE the marginal likelihood (P(Data|M_i_) of each model was estimated through naive monte-carlo estimation. From the estimated marginal likelihood, a Bayes factor comparing each of the three models was calculated.

The model comparison revealed that of the three models (Null = -24146 : condition = - 19283: Individual = -19627), the group model was found to have the largest log likelihood. From these estimated marginal likelihoods, a Bayes factor found that the condition model was heavily favored compared to both the null (Bayes Factor > 10) and individual model (Bayes Factor > 10). The results from this model comparison suggest that there are differences between the parameters estimated for the meAIH and SHAM condition, but not enough evidence to suggest unique individual differences between participants within each condition.

### Model Fit

In Figure 11, the average (+/- 95% HDI) of the condition model’s posterior estimates of the participant’s accuracy and response time reaction time for both the first and second experimental sessions are shown. Overall, the PPE provided a good fit for the average performance of the participants’ individual accuracy (*r* = 0.71, *RMSD* = 0.10) and response time measures (*r* = .51, *RMSD* = 0.36).

**Figure 11.**
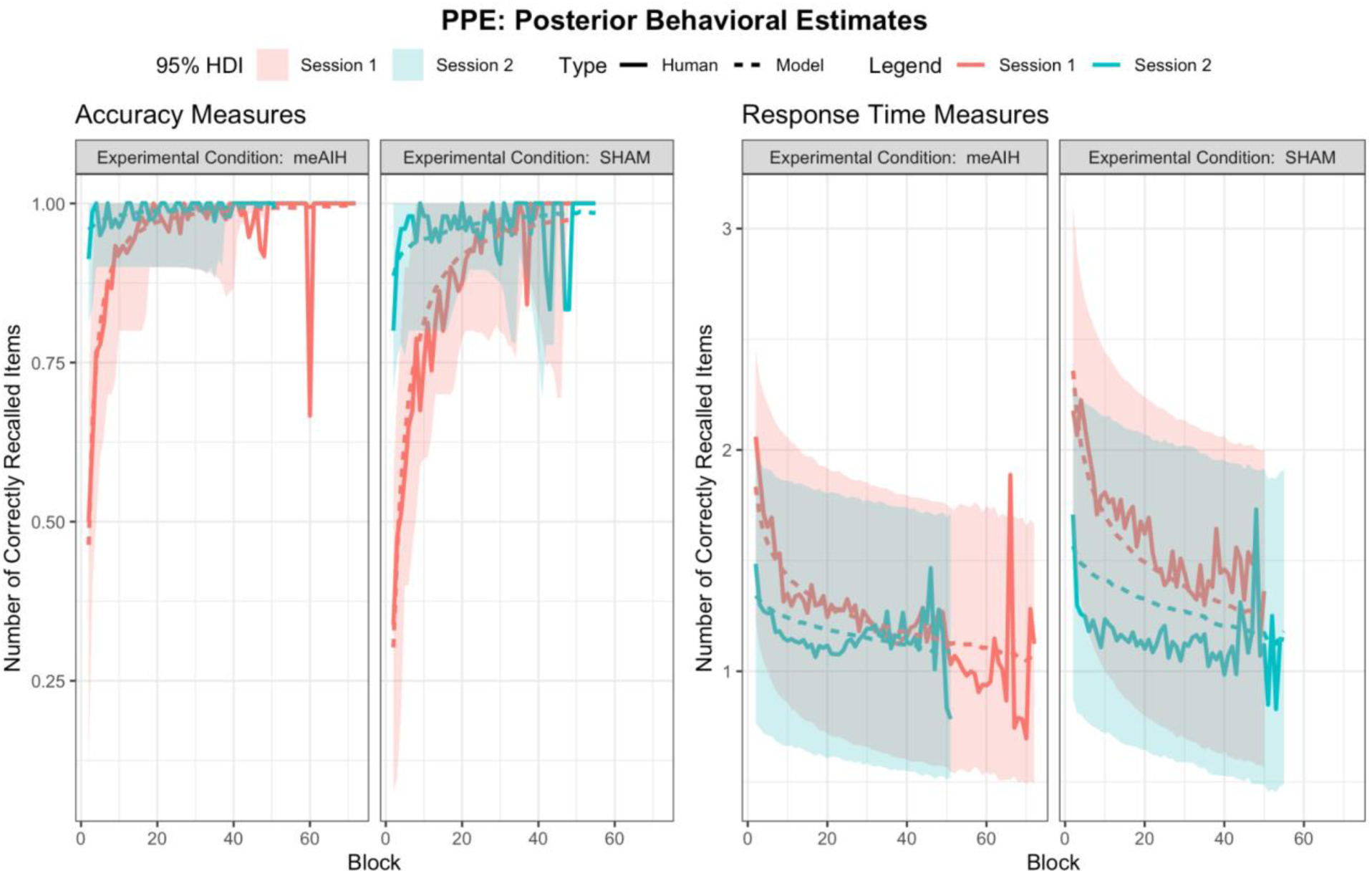
The average accuracy (left plot) and response times (right plot) of both the participants performance (sold line) and estimated posterior (dashed line +/- 95%CI) during the first (red line) and second (blue line) session.

### Parameter Estimates

From the PPE’s fit to the participants’ performance, the estimated posterior parameters from the group model between the meAIH and SHAM condition were then compared (Figure 12). The posterior parameter estimates are a measure of the certainty in a particular parameter value given the model’s prior and observed data. An examination of the posterior distributions between the parameters estimated for the meAIH and SHAM condition revealed particular differences between the two conditions, with some parameters having smaller differences between conditions (b,m,s) compared to others (***τ***, F, f, k).

**Figure 12.**
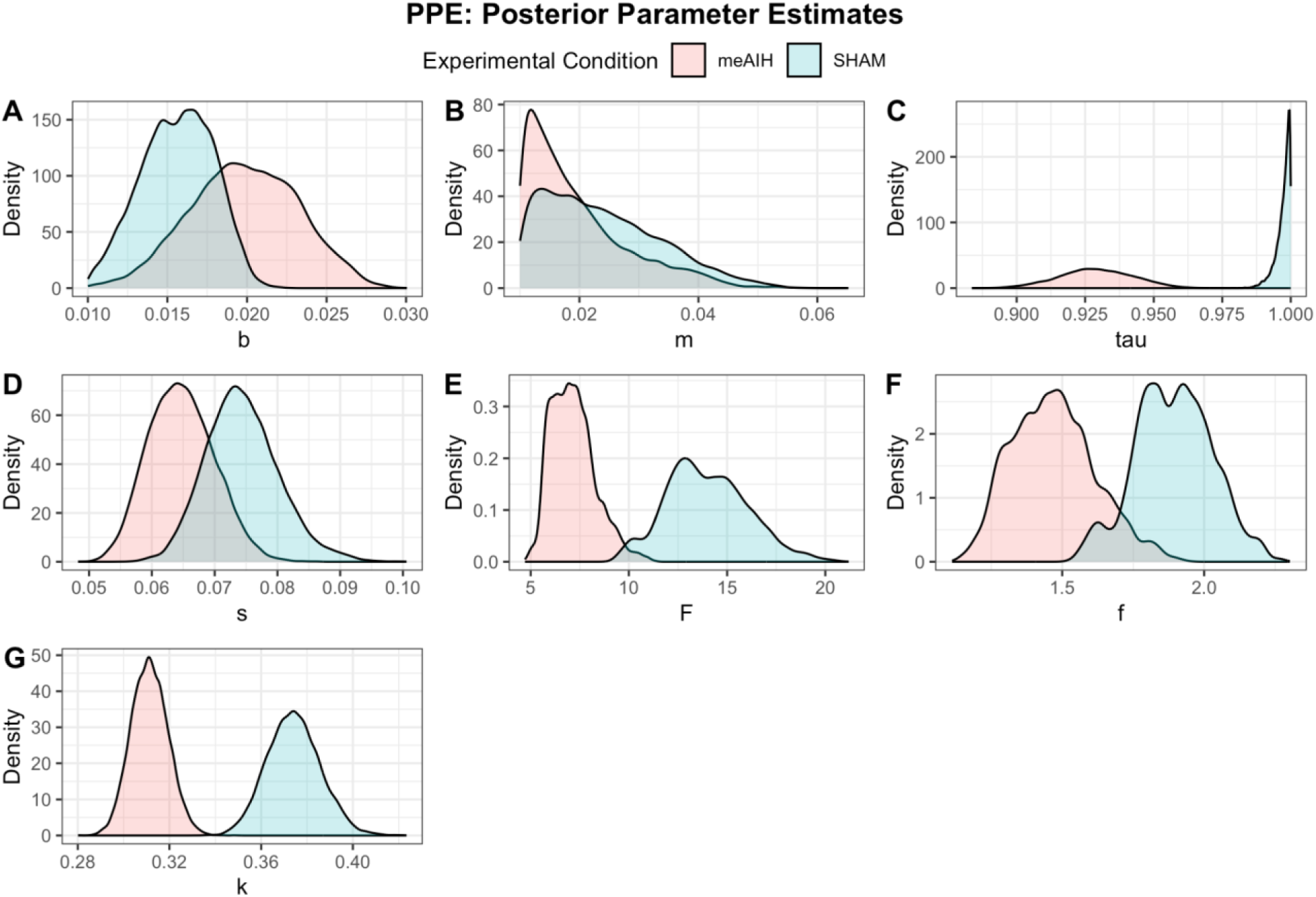
The posterior distribution across the different PPE parameters (b - panel A, m - panel B, tau - panel C, s - panel, D - panel E, f - panel F, k - panel G) for the SHAM (blue) and meAIH (red) condition.

To evaluate the degree of difference in the parameter estimates between the meAIH and SHAM conditions the estimated difference for each estimated parameter was taken (Figure 13). If the 95% HDI (Figure 12 - vertical black lines) between the two estimated parameter values includes 0, then we concluded that there was no significant difference between the estimated parameter between the two conditions. If the 95% HDI between the two estimated parameter values does not include 0, we concluded that there was a significant difference between the estimated parameter between the two conditions. From Figure 12, it can be assumed that there was a significant difference between the ***τ*** (Figure 12 - panel C), F (Figure 12 - panel E), f (Figure 12 -panel F), and k (Figure 12- panel G) parameters between the meAIH and SHAM condition.

**Figure 13.**
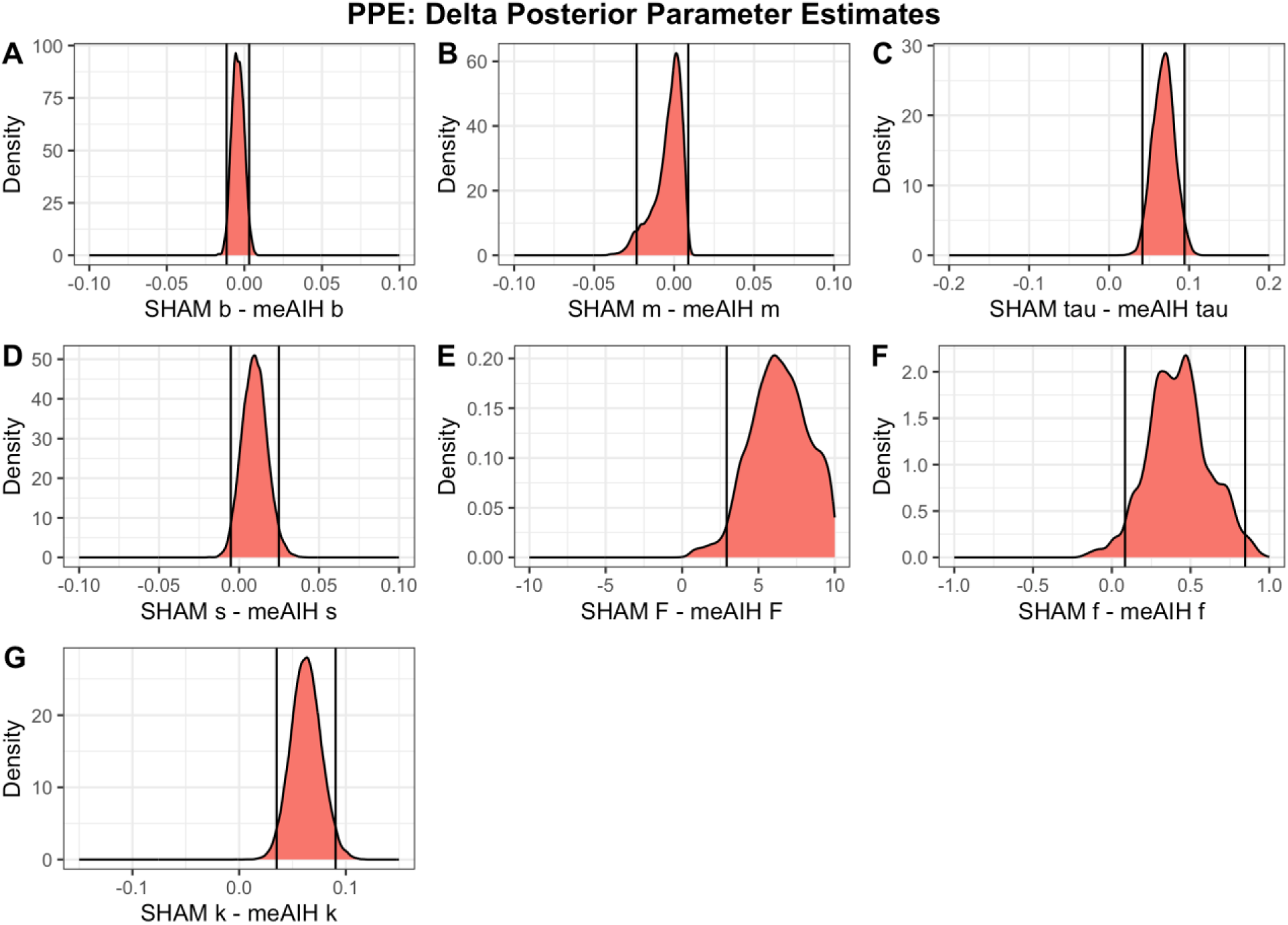
The difference in the posterior distribution between the SHAM and meAIH conditions across each of the PPE parameters (b - panel A, m - panel B, tau - panel C, s - panel, D - panel E, f - panel F, k - panel G). The 95% HDI are marked as straight black lines.

From these differences, we can infer some potential effects of the meAIH manipulation on memory. For the two parameters that affect both accuracy and response times (b and m) no significant difference was found between the conditions, suggesting that the underlying activation quantities for each character-number pair remained the same. The parameters that were found to be different between the two conditions were found to be in the transformation functions for the accuracy and response times. For accuracy we found that the tau parameter was significantly lower in the meAIH condition compared to the SHAM condition. The tau parameter represents the activation threshold for recall, with a lower threshold allowing memories with a lower activation value to have a higher probability of being recalled. The difference in activation thresholds of participants in the meAIH and SHAM conditions could explain the initial higher degree of accuracy in the meAIH compared to the SHAM condition during the first experimental session, when the activation for each learned character number pair is lower and increases with additional presentation. As for response times, there was a significant difference between both the intercept (*F*) and slope (*f*) of the response time function, with the meAIH condition having both a lower response time intercept and slope. As with accuracy the difference between the estimated value of these two parameters suggest possible modification of the meAIH manipulation on memory recall, allowing memories with lower activation to be recalled faster compared to the SHAM condition during the first experimental session.

## Discussion

In this study, the effects of an meAIH protocol on declarative and procedural memory tasks were examined in a young, healthy adult population. Two groups, meAIH and SHAM, participated in an acquisition and retention phase of the study. To further characterize observed results between the meAIH and SHAM conditions, the PPE was implemented as a hierarchical model to infer differences in memory constructs between the groups. Results indicated that the meAIH group differed significantly in the paired associate task accuracy, total errors made, and response times compared to the SHAM group during the acquisition phase. Surprisingly, meAIH had a negative impact on CST accuracy. Just as surprising, the meAIH condition produced faster response times. Bayesian hierarchical modeling revealed that some of these differences may be explained by hippocampal learning thresholds. Collectively, these findings provide an initial demonstration of meAIH accelerating and enhancing the acquisition of knowledge when compared to a SHAM group.

### Effects of meAIH on the Paired Associates Task (PAT)

The meAIH group acquired information more quickly and made fewer errors as the trials progressed through the PAT, compared to the SHAM group. Interestingly, these results do not seem to be an effect of a speed-accuracy trade-off (Salthouse, 1979). Further, it has been reported by Donkin and colleagues (Donkin et al., 2014) that the speed-accuracy trade off phenomenon may be exacerbated by redundant-target trials, lending itself susceptible to the effects of learning. Based on the results, it is plausible the neuroplastic effects that have been previously documented in the administration of AIH may have strengthened the declarative memory containing the associated pair (Schega et al., 2013; Wang et al., 2020). Strengthened memories have been shown to result in faster and more reliable retrievals; it appears meAIH provided a ‘boost’ to require approximately half the number of repetitions to reach asymptotic performance relative to SHAM (see Figure 6, Panel B).

A plausible, mechanistic explanation for the enhanced knowledge acquisition in the meAIH group may be related to the elicited upregulation of BDNF within the hippocampal regions; specifically, the CA3 region. The CA3 region has been thought to be directly responsible for mediating novel information, environments, and experiences (Ergorul & Eichenbaum, 2004; Rolls & Treves, 2024). It is reasonable to suspect that the meAIH group demonstrated an accelerated acquisition of novel information, when exposed to meAIH, in accuracy and button responses due to the upregulation of hippocampal substrates, like the CA3 region.

### Effects of meAIH on the Change Signal Task (CST)

The CST was administered to assess the effects of meAIH on an, overall, procedural memory task. Interestingly, meAIH produced faster response times but poorer accuracy. This was unexpected. However, according to Logan and Burkell (1986), CST paradigm performance relies on multiple neural region contributions (i.e., the medial prefrontal cortex and the CA2 projections to the CA3 regions of the hippocampus). The medial prefrontal cortex interacts with the CA2/CA3 projections from the hippocampus to, ideally, keep information synchronously flowing between these structures (Preston & Eichenbaum, 2013). Perhaps the administration of the meAIH protocol is responsible for the increased response times across tasks at the cost of accuracy in the CST. As a result, meAIH artificially impedes participants ability to inhibit their CST response until they are confident a change-signal was not going to occur. More research is needed to better understand this consequence of meAIH.

### PPE Hierarchical Model

The results from the parameter estimates of the PPE and the participants’ PAT performance revealed two important findings. First, there was a significant difference between the parameters between the meAIH and SHAM condition, but not enough evidence to suggest individual differences across participants. Second, a comparison of the PPE’s estimated parameters between the meAIH and SHAM condition provided potential insights into the effects that meAIH had on memory mechanisms. No significant difference was found in the shared parameters that affect both accuracy and RT measure (*b* and *m*), suggesting that the underlying activation value (*M_i_*) for memories in the meAIH and SHAM condition were not different.

However, parameters related to the transformation function for both accuracy (*τ*) and response times (*F*, *f*), were found to be significantly lower in the meAIH versus the SHAM condition.

One interpretation of the differences in the estimated parameters is that the meAIH manipulation lowered the activation threshold for memories, allowing for a higher probability and faster recall of an item. This interpretation is consistent with Bjork and Bjork (1992) and Walsh et al., (2018) that memory performance (i.e., accuracy and recall) is affected by two separate components (1) storage (e.g., activation) and (2) recall (e.g., transformation function) strength.

Furthermore, a plausible explanation of neuroplastic upregulation can be attributed to changes elicited by BDNF and TrKB (Navarrete-Opazo and Mitchell, 2014; Vose et al., 2022). Similar results have been observed in elderly, mildly cognitive impaired human population (Schega et al., 2013). Participants were administered an AIH protocol, paired with aerobic exercise. Researchers reported improved performance on the six-minute walk test, the California verbal test, and the Mini Mental Status Exam; indicating that an AIH protocol may result in improved cognitive performance. With only the preliminary findings from this paper, strong conclusions of the exact mechanistic effects of meAIH should be avoided, but results appear to be in line with previous research and point to plausible theoretical and physiological explanations of the observed results.

### Limitations and Future Directions

When administering meAIH to a group of young, healthy adults, significant differences were observed in cognitive performance, specifically accuracy and response times during the acquisition time point, when compared to a SHAM group. While these findings are exciting, there are limitations to be discussed. The current study demonstrated proof of concept; however, certain physiological measures were not obtained. For example, O2 saturation, End Tidal CO2, airflow, pressure, or heart rate were not digitally collected during the study. In future studies, surface electromyography coupled with neural measures would allow for a more thorough analysis of neurophysiological state during protocol administration.

Regarding the PAT, three limitations should be addressed. First, during the second experimental session participants were given an initial training trial for each character number pair. Effectively, this served as a refresher training for participants, meaning their ability to recall items after a week-long lag was obscured. The effect of this is that potential differences in the effects of meAIH on long term recall were masked. Future research should keep training trials to only an item’s first presentation during the first experimental session when participants have no knowledge of the task. Second, the learning schedules of the individual PAT items were randomized over the course of an experimental session. The effect of this procedure is that participants were exposed to some items more than others providing unbalanced practice across all items in the PAT. Third, though this procedure was randomized across participants and participants quickly reached ceiling across all items, it does weaken the measure of memory recall over time. Future research should implement additional controls to balance the number of presentations across all items in the PAT.

Finally, the effects observed in this experiment, though significant, ranged from small to very small. Future research should attempt increasing the observed effect size of memory performance between an meAIH and SHAM condition by modifying the PAT task. Over the course of the experiment participants received a great amount of practice across items. A potential effect of this much practice could be that participants were over trained on the PAT items. Future research should attempt to further maximize potential differences by (1) increasing the difficulty of the PAT items or (2) decreasing the number of practice attempts and increasing the lag between individual item presentations.

Item difficulty could be manipulated by modifying the PAT stimuli from number and character pairs to pairs of English words and a secondary language (e.g., Japanese). Decreasing the number of practice items could be accomplished by increasing the number of items on the PAT task and having multiple items on different learning schedules. Items can be set on a learning schedule where they are presented a fewer number of times with longer lags in between presentations, making learning more difficult. While other items can be used as essentially “filler” items presented more frequently and are not the focus of the study. These two manipulations to the PAT task would potentially yield results of slowing the rate of learning, allowing for larger differences between the meAIH and SHAM condition to be observed.

## Conclusion

Importantly, both groups demonstrated learning effects at each time point. This indicates that the paired associate task did elicit learning effects in a young, healthy adult population regardless of the group assignment. Study results seem to indicate that the group that received a calibrated 10.2% oxygen concentration received greater cognitive benefit than those participants who received a calibrated 21% oxygen concentration. Further, the meAIH perturbation demonstrated fewer errors than the SHAM group at acquisition. These results were validated by a PPE hierarchical model. Further, while the results from this research provide a promising framework for increasing learning rates with a well-tolerated and easily administered stimulus, replication of meAIH’s beneficial effects is warranted.

## Data Availability

All raw data files, and code, are available in a repository (see https://drive.google.com/drive/folders/11Ql1TjQiVdsYm7FQN8543-Pda-EUJHh_).

## Footnotes

1 The constant learning rate of .1 is a historical aspect of the PPE coming from its development see Walsh et al. (2018) for details.

